# Patient-Specific Haemodynamic Analysis of Virtual Grafting Strategies in Type-B Aortic Dissection: Impact of Compliance Mismatch

**DOI:** 10.1101/2023.06.20.23288122

**Authors:** Louis Girardin, Catriona Stokes, Myat Soe Thet, Aung Ye Oo, Stavroula Balabani, Vanessa Díaz-Zuccarini

## Abstract

**Introduction:** Compliance mismatch between the aortic wall and Dacron Grafts is a clinical problem concerning aortic haemodynamics and morphological degeneration. The aortic stiffness introduced by grafts can lead to an increased left ventricular (LV) afterload. This study quantifies the impact of compliance mismatch by virtually testing different Type-B aortic dissection (TBAD) surgical grafting strategies in patient-specific, compliant computational fluid dynamics (CFD) simulations.

**Materials and Methods:** A post-operative case of TBAD was segmented from computed tomography angiography data. Three virtual surgeries were generated using different grafts; two additional cases with compliant grafts were assessed. Compliant CFD simulations were performed using a patient-specific inlet flow rate and three-element Windkessel outlet boundary conditions informed by 2D-Flow Magnetic Resonance Imaging (2DMRI) data. The wall compliance was calibrated using Cine-MRI images. Pressure, wall shear stress (WSS) indices and energy loss (EL) were computed.

**Results:** Increased aortic stiffness and longer grafts increased aortic pressure and EL. Implementing a compliant graft matching the aortic compliance of the patient reduced the pulse pressure by 11% and EL by 4%. The endothelial cell activation potential (ECAP) differed the most within the aneurysm, where the maximum percentage difference between the reference case and the mid (MDA) and complete (CDA) descending aorta replacements increased by 16% and 20%, respectively.

**Conclusion:** This study highlights the negative impact of increased graft length on LV condition after surgical aortic replacement in TBAD. To mitigate the associated risks to the patient, graft manufacturers should allocate more resources toward developing compliant biomimetic grafts.

## Introduction

Type-B Aortic Dissection (TBAD) is a cardiovascular disease involving a tear in the descending aorta where the dissected portion can extend into the abdomen and legs. In 91% of chronic TBADs, patients will survive the initial dissection, and 60% of them will develop late aneurysmal dilation of the dissected portion, organ, or limb malperfusion, which may require surgical treatment in 25-50% of cases [1][2]. The relative merits of Thoracic endovascular aortic repair (TEVAR) and open surgery (OS) have been widely debated [3]. TEVAR has better early outcomes than OS, but TEVAR had significantly worse long-term survival rates compared to OS in a study of over 15,000 patients [4]. TEVAR is increasingly favoured over OS in cases of chronic dissection due to higher mortality risk associated with OS. In a systematic review of 19 studies, it was found that the overall mortality rate for type B aortic dissection open surgery is 11.1%, which is reduced to 7.5% in the endovascular era [5]. However, OS is necessary in about 30% of cases when the chronic dissection is complicated and not amenable to endovascular treatment or in the case of connective tissue disorders that compromise graft landing zones [6]. Indeed, complications such as pulmonary collapse, renal failure or malperfusion leading to ischemia may develop in the acute or chronic stages, requiring immediate surgery. During OS, the aorta is clamped, cut, and replaced with a Dacron Graft to reconstruct the aortic blood flow through thoraco-phreno-laparotomy. Unfortunately, OS has been associated with poor outcomes [7], long-term adverse effects such as retrograde left ventricular (LV) hypertrophy and the development of new antegrade aortic tears in the aortic wall [8].

The compliance mismatch between the rigid graft and the remaining aorta can be detrimental as it can increase aortic impedance and pressure [9][10]. Aortic impedance refers to the resistance to blood flow by the aorta and is believed to cause increasing aortic diameter and stiffening of the vessel, which can increase the afterload and the amount of resistance the heart must overcome to pump blood out of the LV to the aorta. The rigid graft introduces an impedance mismatch with the aorta due to their different elastic properties. Reflections of pulse waves may be expected, which can lead to pressure increase [11][12]. Increasing aortic impedance can lead to alterations in blood flow and increased energy loss (EL), as blood must flow through sections of the aorta that offer more resistance to flow [13]. EL, defined as the comparison of how much energy is used during the stretching of the aorta in diastole and the relaxing of the stored energy in systole [14], subsequently increases with the rigid graft, leading to greater stress on the heart, and additional LV mass, ultimately resulting in hypertrophy [15].

Furthermore, the compliance mismatch reduces the total aortic compliance, altering the ability of the aorta to expand during systole in response to higher blood volume and pressure. Decreasing aortic compliance is another risk factor for LV hypertrophy [16]. In addition, the increase in aortic impedance and decrease in aortic compliance brought by the graft causes pressure waves to travel faster through the aorta and increase pulse wave velocity (PWV) [17]. An increasing PWV has been associated with an increased risk of cardiovascular events such as myocardial infarction and stroke [18]. The choice of graft length and diameter is a critical choice for surgeons as the deterioration of cardiac function is closely linked with aortic stiffness. Clinicians must carefully balance graft length and aortic rigidity to lead to a favourable outcome.

Blood pressure is currently the standard metric used in clinical practice as high blood pressure (i.e., >140 mmHg) have been linked with increasing risk of AD [19]. Other metrics such as velocity, aortic wall displacement can be derived from MRI images. While they are not yet of standard use, these metrics have the potential to guide surgical decision-making by more directly influencing physiological mechanisms than existing anatomical predictive metrics. Wall shear stress (WSS) and pressure distributions have been shown to affect the homeostatic regulation of vessel wall structure and are linked to aortic wall degeneration [2][20]. Thus, precise quantification of these metrics may help predict wall degeneration and unfavourable disease progression. Non-invasive medical imaging techniques such as 2D-Flow Magnetic Resonance Imaging (2DMRI) and 4D-Flow MRI (4DMRI) can measure aortic blood velocity, while Cine-MRI can measure wall displacement [20]. However, the metal coil within grafts can produce image noise during acquisition, and MRI scanners are not available in all clinical settings [21][22]. Furthermore, 2DMRI and 4DMRI are subject to numerous errors, cannot measure pressure distributions and fail to accurately capture WSS [23]. Computational fluid dynamics (CFD) can enhance in vivo imaging by accurately calculating relevent haemodynamic metric. This is even more relevant in the case of limited clinical datasets. CFD allows for higher spatial and temporal resolution than most imaging techniques [24][25], and can enable virtual testing of various surgical techniques and sizing of medical devices, which is impossible to do during surgery [26][27].

Although CFD has shown potential to help understanding disease mechanisms, it is not yet suitable for treating patients with TBAD. This is due to a lack of large-scale clinical studies and standardised simulation practices.

There are several challenges in employing CFD for aortic dissection (AD) intervention planning. AD is complex and highly patient specific. The aortic wall is compliant, i.e., it can stretch and contract with the pulsatile flow of blood [28] while devices such as grafts are rigid which may cause mechanical deformation of the aortic wall, affecting the blood flow dynamics and leading to tissue damage or rupture [8]. Therefore, robust flow modelling techniques that account for aortic wall compliance are necessary to accurately capture the interaction of surgical devices such as grafts with the aortic wall and the impact of aortic compliance in a patient-specific manner [29][30]. While the assumption of rigid wall can provide a simple and fast approach to virtual surgical intervention planning, particularly in cases where AD wall movement is minimal, compliant simulations have been found to yield more accurate predictions of WSS calculations than rigid wall simulations [31][32]. The stiffness of the aortic wall and intimal flap are important factors that affect haemodynamics and WSS distribution in TBAD [33][34]. Consequently, to investigate the effect of TBAD OS on LV afterload, it is necessary to use a compliant simulation method that can accurately model the interaction between the grafts, aortic wall, and blood flow [17].

Fluid-structure interaction (FSI) is commonly employed to account for wall compliance in patient specific cardiovascular modelling. An FSI study of aortic valve sparing reconstruction [35] were shown to correctly capture the haemodynamic effects induced by the compliance mismatch between the stiff grfat an dthe native vessel. FSI has been also applied in the context of TBAD to study the effect of endograft length on LV; a simplified geometry and literature-based boundary conditions were employed [36]. The LV workload was reduced when using a medium-length endograft; the latter also resulted in false lumen flow reversal which is thought to induce false lumen thrombosis. However, FSI is associated with various limitations, such as its dependence on assumptions about the mechanical properties of the vessel wall, which are patient-specific and cannot be directly measured *in vivo*. It also requires substantial computational resources and is particularly challenging to implement. The Moving Boundary Method (MBM) [33], is an efficient alternative to FSI, eliminating the need for structural modelling of the aortic wall and the associated material assumptions. The MBM is less computationally demanding than FSI, whilst capable of providing the same level of accuracy in mimicking aortic wall displacement [33].

In the present study we employ the MBM [33] method to account for graft and aortic wall compliance in patient-specific CFD simulations to provide insights into the clinical significance of graft length and aortic compliance mismatch in the context of OS for TBAD, which has been limited to date. CFD simulations are informed by a set of routinely obtained medical imaging data, including, computed tomography angiography (CTA) and limited time-resolved Cine-MRI and 2DMRI data and used to explore a number of surgical graft scenarios and their impact on blood flow distribution and LV afterload both proximal and distal to the graft site.

## Materials and Methods

### Data acquisition

A patient with a complicated chronic TBAD was presented at St Bartholomew’s Hospital, London, UK. The patient underwent OS where a dissected portion of the thoracic aorta was replaced with a graft (Gealweave, Terumo Aortic, Vascutek LTD, UK). The graft was 130 mm long with a 32 mm diameter (see Fig. 1A). Their aorta was imaged prior to and after OS. Following an ethically approved protocol (St Bartholomew’s Hospital BioResource ethical application number 97), Cine-MRI and 2DMRI were acquired pre-operatively using a Siemens MAGNETOM Aera 1.5T (Siemens Healthcare GmbH, Erlangen, Germany) with a resolution of 1.7 mm*1.7mm. 2DMRI were acquired at one plane 5 cm distal to the primary entry tear (PET), located 36 mm distal to the aortic arch. CTA images were also acquired as part of the routine post-operative clinical examination (see Fig. 1A) using a Siemens SOMATOM Definition Edge with a resolution of 0.73 mm*0.73 mm*0.75 mm. Brachial pressures were acquired post-operatively. It should be noted that the patient was on medication with beta-blockers to reduce arterial pressure.

**FIGURE 1.**
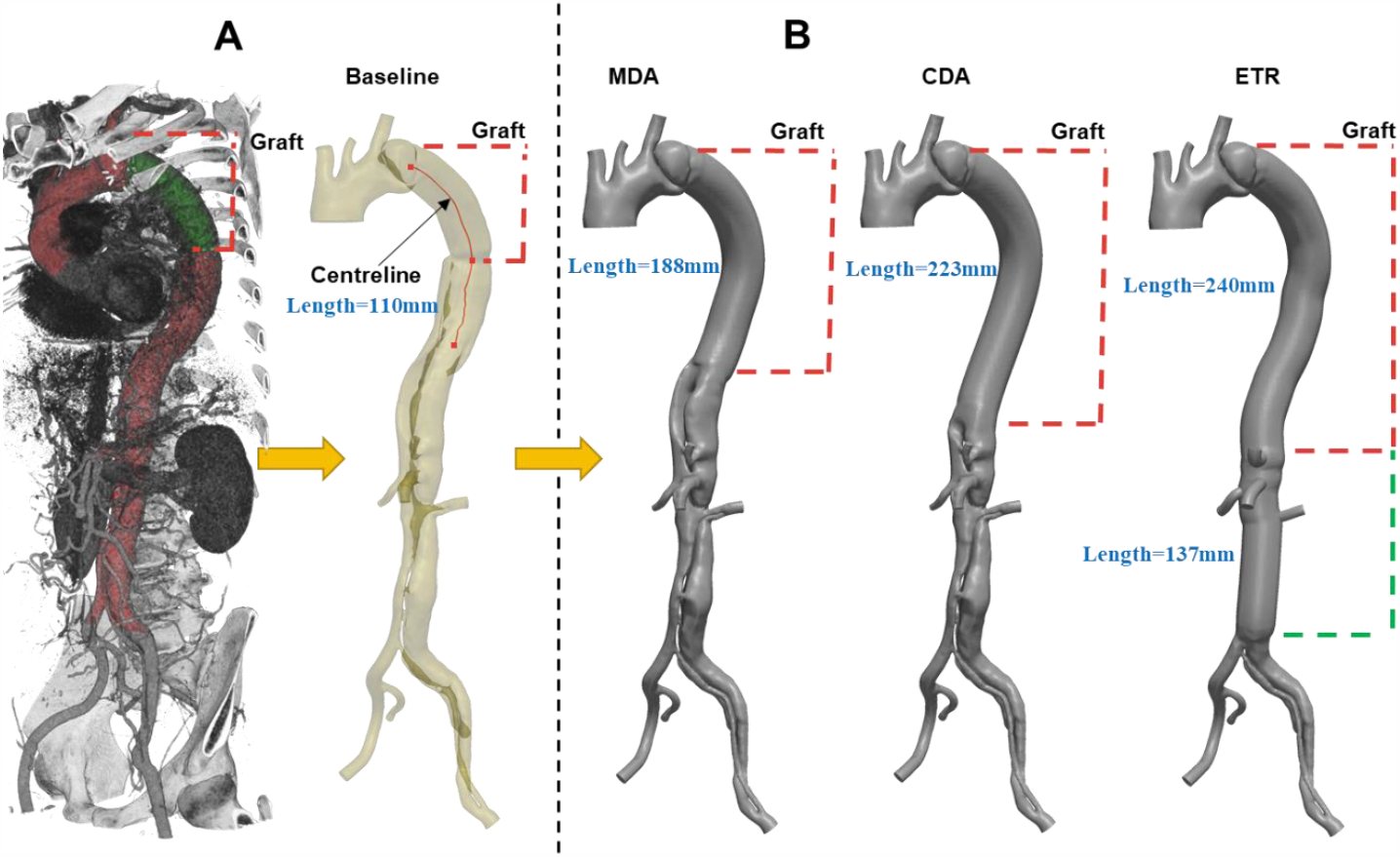
A Automatic 3D rendering of the CTA, with, in red, the aortic vessel and, in green, the graft; next is the segmented DA resulting from the CTA. B The three virtual surgeries created from the baseline case by varying the length of graft. The red centreline from which the grafts have been swept is shown on the post-operative geometry. Red and green dashed lines indicate the extent of the 32mm graft and 28mm thoracoabdominal graft of ETR, respectively. The length of each graft is in blue.

### Image processing and Virtual Surgical Interventions

The CTA data (Fig. 1A) were segmented using automatic thresholding and manual correction of the mask implemented in ScanIP (Synopsis Simpleware, USA). The clinical team verified the segmented geometry, confirming the location of the tears. The resulting mask was then smoothed using Meshmixer (Autodesk, USA). The inlet and all outlets, namely the brachiocephalic trunk (BT), left common carotid (LCC), left subclavian (LSA), celiac trunk (CT), superior mesenteric artery (SMA), right (RR) and left renal (LR), left exterior (LEI) and interior iliac (LII), right exterior (REI) and interior iliac (RII), were trimmed so that their cross-sectional areas were perpendicular to the flow direction using Fluent Mesh (Ansys Fluent, USA) (Fig. 1B). Three virtual grafting scenarios were subsequently created in consultation with the clinal team, by extending the graft using ScanIP and Meshmixer. Two lengths corresponding to the descending half and total length of the aorta were considered, denoted as mid-descending (MDA) and complete descending (CDA) aorta, respectively. The third grafting scenario involved an entire replacement of the thoracoabdominal aorta (ETR) to the iliac bifurcation. Two additional cases with compliant graft were simulated using the baseline case geometry. The first was named baseline compliant 1 (BC1), the second case, named BC2+. The two cases are described further down in the section “simulation of wall dispalcement and compliant graft cases”.

### Computational Mesh

Tetrahedral computational meshes were created for each domain using Fluent Mesh 19.0 (Ansys Inc., USA). Maximum and minimum cell sizes were identical across cases (4 mm, 0.35 mm). Ten prism layers with a first layer corresponding to a y+ of 1 were used to ensure appropriate boundary layer modelling for each mesh. A mesh independence study was conducted using the baseline case; coarse, medium, and fine meshes were generated by approximately doubling and dividing the maximum and minimum element sizes. The Grid Convergence Index, detailed by Craven et al. [37], was used to assess the quality of the baseline mesh. The index did not exceed 4.5% on every mesh for all metrics, consistent with past research [37]. More details are available in the supplementary materials. Using the final mesh resolution determined from the mesh independence study, the baseline, MDA, CDA and ETR cases contained 1.35, 1.2, 1.1 and 0.9 million elements, respectively.

### Boundary Conditions

The inlet flow rate was extracted from the pre-operative 2DMRI data near the aortic arch using GTFlow (GyroTools LLC., Switzerland) (Fig 2A). It has been reported that approximately 30% of the blood leaves the aorta through the supra-aortic branches supra-aortic branches [38]. Hence, the measured flow rate at the arch was scaled by 30% to represent the inlet flow rate. The flow rate curve was spline-interpolated in MATLAB (MathWorks Inc., USA) to match the CFD timestep of 1 ms and was used to apply a uniform inlet velocity profile (Fig 2A).

**Figure 2.**
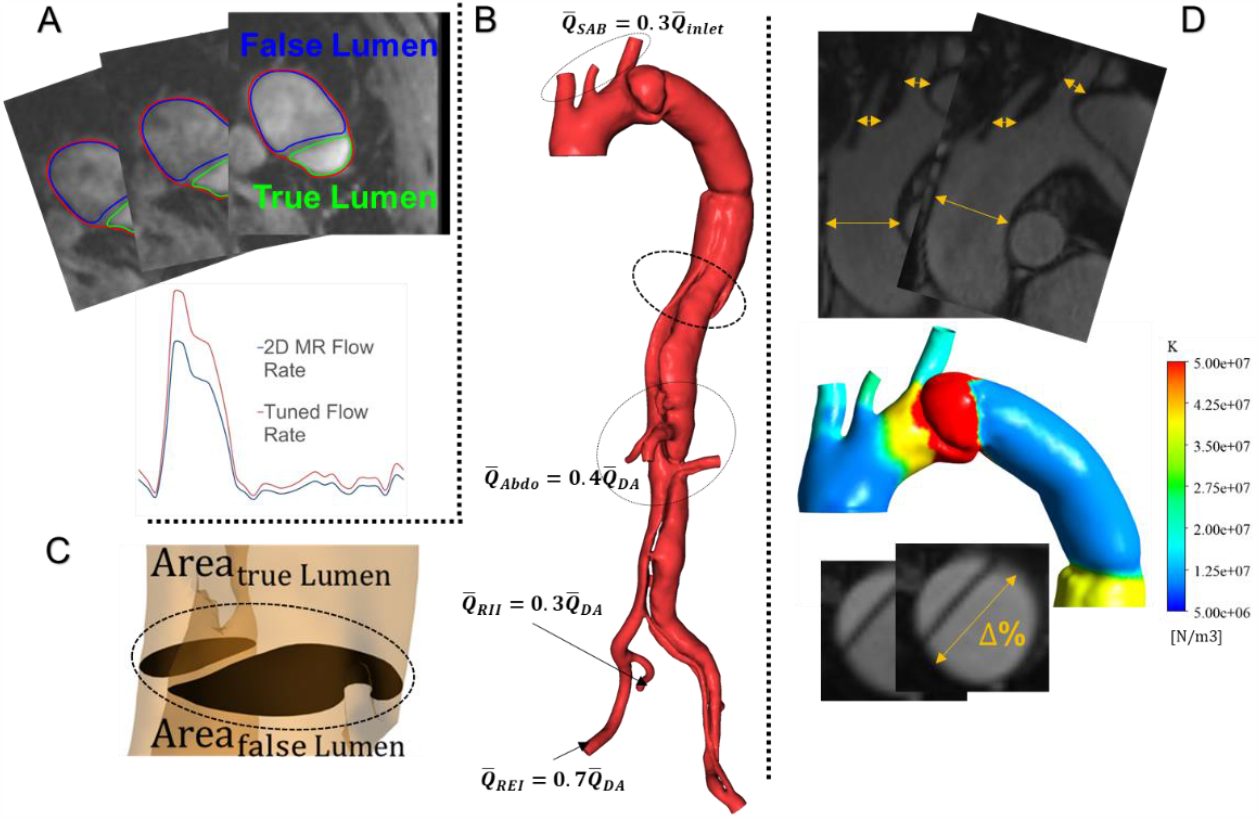
A 2DMRI plane showing in blue and green the false and true lumen respectively; below are the extracted raw and rescaled by 30% flow rates, B Flow split at the outlets: 30% of the flow leaves through the supra-aortic branches, and 40% of the remaining DA flow leaves through the abdominal arteries following the true and false lumen shown in C. The remaining abdominal false and true lumen flows are split as 70/30% between the exterior and interior iliac arteries; the right exterior (REI) and interior (RII) iliac arteries are shown as an example. D Sample cine-MRI planes used to measure the stiffness of the aorta. The aortic arch of BC1 is zoomed in to show the distribution of local stiffness values K obtained for the case of a compliant graft.

Three-element Windkessel (WK3) outlet pressure boundary conditions were applied to mimic the effects of the peripheral vascular system. WK3 are electrical analogues in which a proximal resistance *R*_*p*_, a distal resistance *R*_*d*_ and a capacitance *C*_*WKS*_ are used to the haemodynamic behaviour of the distal vasculature. The systolic pressure *P*_*sys,a*_, diastolic pressure *P*_*dia,a*_ and the mean flow rates at the outlets are required to calibrate the WK3 elements. These were obtained from brachial measurements (*P*_*sys,b*_, *P*_*dia,b*_) were used to calculate *P*_*sys,a*_ and *P*_*dia,a*_. Following Westerhof et al. [28], the diastolic pressure was taken to be constant in the arterial tree, such that *P*_*dia,a*_ = *P*_*dia,b*_, and the systolic aortic pressure was set to be equal to *P*_*sys,a*_ = 0.83 *P*_*sys,b*_ + 0.15 *P*_*dia,b*_. Target mean flow rates at each outlet were split as follows: 30% of the flow was assigned to the supra-aortic branches, and the mean flow rates for each branch were calculated by dividing the total supra-aortic branches flow by their respective cross-sectional area ratio, such that:

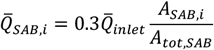

where 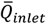is the mean flow rate at the inlet over a cardiac cycle (mL/s), *A*_*SAB,i*_ is the cross-sectional area of the supra-aortic branches outlet (*m*^2^), and *A*_*tot,SAB*_ the sum of the supra-aortic branches cross-sectional area (*m*^2^). The distribution of blood flow in the abdominal region varies among patients and can be affected by the precise nature of the dissection. A study by Amanuma et al. [39] found that the blood flow leaving the abdominal branches ranged from 25% to 75% in a group of 10 patients. After consultation with the medical team, the mean flow leaving the abdominal arteries was set as 40% of the residual flow in the descending aorta after OS. The abdominal branches are perfused by both lumens, as shown in Figure 2B-C. Hence, the mean flow rates to the abdominal branches were determined using a cross-sectional area split method, such that:

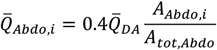

where 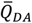 and 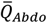 are the descending aorta and abdominal branches mean flow rates over a cardiac cycle (mL/s), *A*_*Abdo,i*_ is the cross-sectional area of the abdominal branches outlet (*m*^2^), and *A*_*tot,Abdo*_ the sum of the abdominal branches cross-sectional area (*m*^2^). The remaining mean flow was split using a 70/30% balance between the external and internal iliac arteries based on the work of Bonfanti et al. [40], as shown in Figure 2B. The same flow split methodology was applied to the four geometries and is summarised in Table 1.

**Table 1.** Mean flow rates at the outlets for each case. Flow splits are very close between the post-operative, MDA and CDA cases due to close morphological similarities. Differences are found in the abdominal and iliac arteries of the ETR case due to idealised abdominal branches of the graft.

| $\bar{Q}_{mean}$ [mL/s] | Baseline | MDA | CDA | ETR |
| --- | --- | --- | --- | --- |
| BT | 18.5 | 18.5 | 18.5 | 18.5 |
| LCC | 5.1 | 5.1 | 5.1 | 5.1 |
| LSA | 12.1 | 12.1 | 12.1 | 12.1 |
| CT | 15.0 | 15.0 | 14.9 | 6.5 |
| SMA | 28.4 | 28.5 | 27.2 | 24.2 |
| LR | 12.2 | 12.2 | 11.6 | 11.6 |
| RR | 6.7 | 6.6 | 7.9 | 9.8 |
| LEI | 2.9 | 2.8 | 3.4 | 4.5 |
| LII | 2.5 | 2.5 | 2.7 | 6.5 |
| REI | 10.2 | 10.2 | 10.1 | 10.2 |
| RII | 5.7 | 5.7 | 5.7 | 10.2 |

Calibration of the boundary conditions using an analogue 0D model was performed to obtain the WK3 parameters following the work of Bonfanti et al. [40] and Stokes et al. [41]. The WK3 parameters obtained after calibration for the post-operative, MDA, CDA and ETR cases are presented in Table 2. The BC1 and BC2+ cases are not included in the table for clarity since the resistances are the same as those of the baseline case where the same flow split is applied to the same geometry.

**Table 2.** WK3 parameters for the Baseline, MDA, CDA and ETR cases, *R*_*p*_ and *R*_*d*_ are in (*mmHg* ∗ *s*/*mL*), *C*_*WKS*_ is in (*mL*/*mmHg*)

|  | Baseline |  |  | MDA |  |  | CDA |  |  | ETR |  |  |
| --- | --- | --- | --- | --- | --- | --- | --- | --- | --- | --- | --- | --- |
| | $R_p$ | $R_d$ | $C_{WK3}$ | $R_p$ | $R_d$ | $C_{WK3}$ | $R_p$ | $R_d$ | $C_{WK3}$ | $R_p$ | $R_d$ | $C_{WK3}$ |
| BT | 0.24 | 4.0 | 0.32 | 0.24 | 4.0 | 0.31 | 0.24 | 4.0 | 0.30 | 0.24 | 4.0 | 0.28 |
| LCC | 0.85 | 14.38 | 0.09 | 0.85 | 14.38 | 0.09 | 0.85 | 14.38 | 0.08 | 0.85 | 14.38 | 0.08 |
| LSA | 0.36 | 6.1 | 0.21 | 0.36 | 6.1 | 0.21 | 0.36 | 6.1 | 0.20 | 0.36 | 6.1 | 0.19 |
| CT | 0.74 | 12.51 | 0.10 | 0.74 | 12.48 | 0.10 | 0.74 | 12.54 | 0.09 | 0.42 | 7.08 | 0.16 |
| SMA | 0.42 | 7.1 | 0.18 | 0.42 | 7.09 | 0.17 | 0.42 | 7.13 | 0.17 | 0.42 | 7.08 | 0.16 |
| LR | 1.41 | 3.62 | 0.26 | 1.39 | 3.56 | 0.25 | 1.4 | 0.26 | 0.25 | 3.28 | 8.43 | 0.10 |
| RR | 8.45 | 21.74 | 0.04 | 8.54 | 21.95 | 0.04 | 7.95 | 20.44 | 0.04 | 3.28 | 8.44 | 0.10 |
| LEI | 0.15 | 2.48 | 0.50 | 0.15 | 2.48 | 0.48 | 0.15 | 2.61 | 0.45 | 0.17 | 2.94 | 0.37 |
| LII | 0.35 | 5.89 | 0.21 | 0.35 | 5.88 | 0.21 | 0.37 | 6.18 | 0.19 | 0.37 | 6.19 | 0.18 |
| REI | 0.64 | 10.75 | 0.12 | 0.66 | 11.05 | 0.11 | 0.54 | 9.06 | 0.13 | 0.44 | 7.36 | 0.15 |
| RII | 1.49 | 25.05 | 0.05 | 1.51 | 25.53 | 0.05 | 1.25 | 21.11 | 0.06 | 0.94 | 15.85 | 0.07 |

### Simulation of Wall Displacement and Compliant Graft Cases

The MBM developed by Bonfanti et al. [33] was applied to simulate aortic wall compliance. The wall displacement follows the surface node normal and is proportional to the difference between local and external pressures; the proportionnality constant is the stifness coefficient, *K*_*i*_. The displacement of each mesh node is thus calculated as follows:

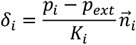

where the local pressure is *p*_*i*_ (Pa), *p*_*ext*_(Pa) is the external pressure (equal to *P*_*dia,a*_). The stifness coefficient *K*_*i*_ (N/m^3^) is equal to:

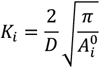

where 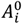 (m^2^) is the local diastolic cross-sectionnal area and *D* (1/Pa) is the local wall distensibility. *D* was calculated using wall movement data extracted from Cine-MRI (Fig 2D) as follows:

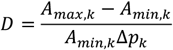

where *A*_*max,k*_ and *A*_*min,k*_ (m^2^) are the maximum and minimum cross-sectional area of the aortic vessel in a given region *k* and ∆*p*_*k*_ is the average pulse pressure in that region, as estimated from a rigid, transient CFD simulation. When axial Cine-MRI images were unavailable, for example at the aortic arch, sagittal images were used to measure wall displacement. The assumption of a circular cross-section in the aorta and supra-aortic branches was employed so that diameters could be used in lieu of the cross-sectional area to calculate distensibility. The distensibility of each region was used to calculate the stiffness coefficient *K*, which was then mapped to its respective region of the geometry using an in-house MATLAB code. As observed in Fig 2D, and following the work of Stokes et al. [41], three smoothing iterations were done to avoid discontinuities between regions of different stiffness. The graft was considered very stiff (K=1.0 10^9^N/m^3^) in the baseline case, MDA, CDA and ETR cases. Two additional cases were simulated. In the first, BC1, the graft was identical to the baseline geometry but the graft stiffness was equal to the measured aortic stiffness at the ascending aorta (*K*_*BC*1_ = 7.5 10^6^N/m^3^) (Fig 2 D). The second case, BC2+, also had an identical geometry to the baseline case but with a graft stiffness two times smaller than BC1 (*K*_*BC*2+_ = 3.75 10^6^N/m^3^). This latter case aimed to simulate a graft which was more compliant than any region of the aorta.

### Computational Model

The three-dimensional, transient Navier-Stokes equations were solved using the finite-volume solver ANSYS CFX 19.0 using the Carreau-Yasuda viscosity model and empirical constants from Tomaiulo et al. [42]. Blood was modelled as an incompressible non-Newtonian fluid with a density of 1056 kg/m3. By using the Reynolds number descriptions for pulsatile blood flow in cardiovascular systems as outlined by Peacock et al. [43], determining the effective shear rate based on the research of Cagney et al. [44], and increasing the maximum velocity from the 2D magnetic resonance (2DMRI) plane by 30% to account for supra-aortic branches flow loss, the peak *Re*_*p*_ and critical *Re*_*c*_ were calculated as 1281, and 5007 respectively. Under these conditions, a laminar flow assumption was used. An implicit, second-order backward Euler scheme with a time step of 1ms was used to solve the Navier Stokes and continuity equations. During the final cycle, all equations within each timestep had a root-mean-square residual value of 10^−5^. After seven cycles, the compliant simulations reached periodic conditions, i.e., less than 1% variation in systolic and diastolic pressures between cycles. Simulations were run on the high-performance computing cluster of the UCL Computer Science Department (computational time: 23h/cycle).

### Haemodynamic Parameters

EL and WSS-driven indices were calculated in this work. EL describes the Windkessel function of the aorta, which stores blood and elastic energy due to aortic compliance during systole and releases back the energy as fluid momentum during diastole. EL is a measurement of the amount of energy that is lost as heat and other energy forms in this process, and thus the amount of energy that is not returned as fluid momentum [14]. EL is related to pressure and flow rate within the aorta. As a result, EL often increases in the case of AD due to increased blood pressure [45]. The heart must work harder to compensate for the increased pressure, EL, and reduced blood flow, potentially leading to heart failure [46]. The rigidity introduced by the graft can also increase EL, possibly contributing to poorer outcomes after OS [8].

The EL is calculated from the difference in the sum of static and dynamic pressures between the inlet and outlets during a cardiac cycle and is defined as follows [47]:

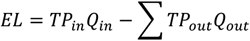

where 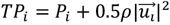 is the blood density (kg/m^3^), *Q*_*i*_ the volume flow rate (m^3^/s), *P*_*i*_ the pressure (Pa), and 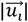 the velocity magnitude (m/s).

Measured as the shear force applied to the inner surface of the arteries divided by area, WSS has been linked to the development of aortic disease [48]. WSS-related metrics usually quantify three indices: time average wall shear stress (TAWSS), oscillatory shear index (OSI), and endothelial cell activation potential (ECAP) [49]. TAWSS is the averaged WSS over a cardiac cycle and measures the total shear stress applied to the wall. OSI measures the axial directional changes of the WSS vector over the cardiac cycle. By definition, OSI varies between 0 to 0.5, indicating unidirectional WSS vector for low values and a fluctuating WSS vector for high values. ECAP is defined as the ratio of OSI and TAWSS and quantifies the degree of thrombogenic susceptibility of the aortic wall. High values of ECAP (>1.4 Pa-1) correspond to regions where the OSI is high and the TAWSS is small, which indicates regions susceptible to high endothelial cell deposition and thrombosis [50]. These WSS indices are as follows [51]:

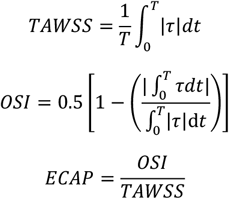

where *T* is the cardiac cycle period (s), and τthe instanteneaous WSS vector.

To better elucidate the impact of the various grafts on hemodynamics, the TAWSS and ECAP differences between the baseline and the five cases examined are computed. The latter are normalized by the baseline average as:

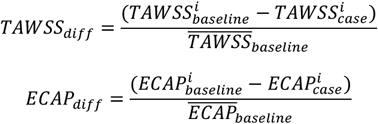

## Results

### Comparisons Between Base Case CFD Results and Targeted Clinical Data

Validation was performed via qualitative and quantitative comparisons between the CFD simulations of the baseline case and the target values from clinical data. The relative error in metrics of interest was calculated and shown in Table 3.

**Table 3.** Systolic *P*_*sys,a*_ and diastolic *P*_*dia,a*_ pressures, mean flow rate *Q*_*mean*_ at the registered plane location and maximum diameter variation at regions of interest for the baseline simulation and clinical data measurements.

|  |  |  |  | Diameter Variation (mm) |  |  |  |
| --- | --- | --- | --- | --- | --- | --- | --- |
| | Psys (mmHg) | Pdia (mmHg) | $Q_{mean}(\frac{mL}{s})$ | AA | BT | LCC | LSA |
| Measured | 96.5 | 68 | 83.5 | 1.3 | 0.7 | 0.5 | 0.5 |
| Baseline | 97.5 | 68.4 | 84.9 | 1.26 | 0.69 | 0.49 | 0.49 |
| Error | -1.0% | -0.6% | -3.2% | 1.4% | 2.0% | 2.0% | -1.0% |

*P*_*sys,a*_ and *P*_*dia,a*_ were obtained within 1% of relative error against the brachial pressure cuff measurements. The simulated aortic wall displacement was compared against the Cine-MRI measurements; the maximum diameter variation over a cardiac cycle was measured. Measurements were taken at the AA and supra-aortic branches where most displacement occurs (Fig 2D); relative errors between the Cine-MRI and baseline measurements were under 2%. The coordinates of the 2DMRI plane were extracted and registered onto the CFD domain to compare the mean flow at the same location. The relative error between the mean flow rates was 3.2% (Fig 2A). As errors remained minor (i.e., 3.2%) between the CFD simulation and the clinical data measurements, the simulation settings were deemed suitable to be applied to the additional intervention cases.

### Pressure, Wall Displacement and Energy Loss (EL)

The LV pressure has been reported to vary linearly with AA pressure [52]; if the AA pressure increases, so does the LV pressure. We report the *P*_*sys,a*_ and *P*_*dia,a*_ at the inlet of the baseline case (Fig 3A) and the relative error with respect to the five additional cases to show the impact of graft length and compliance in pressure values (Fig 3B). Inlet pressure increased with longer grafts. The ETR case had the highest systolic and diastolic pressures, however, the increasing pressure trend with longer graft was not followed, as there was no significant pressure increase compared to CDA. The compliant graft used in BC1 reduced the inlet pressure; however, the trend was not linear as the pressure of BC2+ increased compared to BC1.

**Fig 3.**
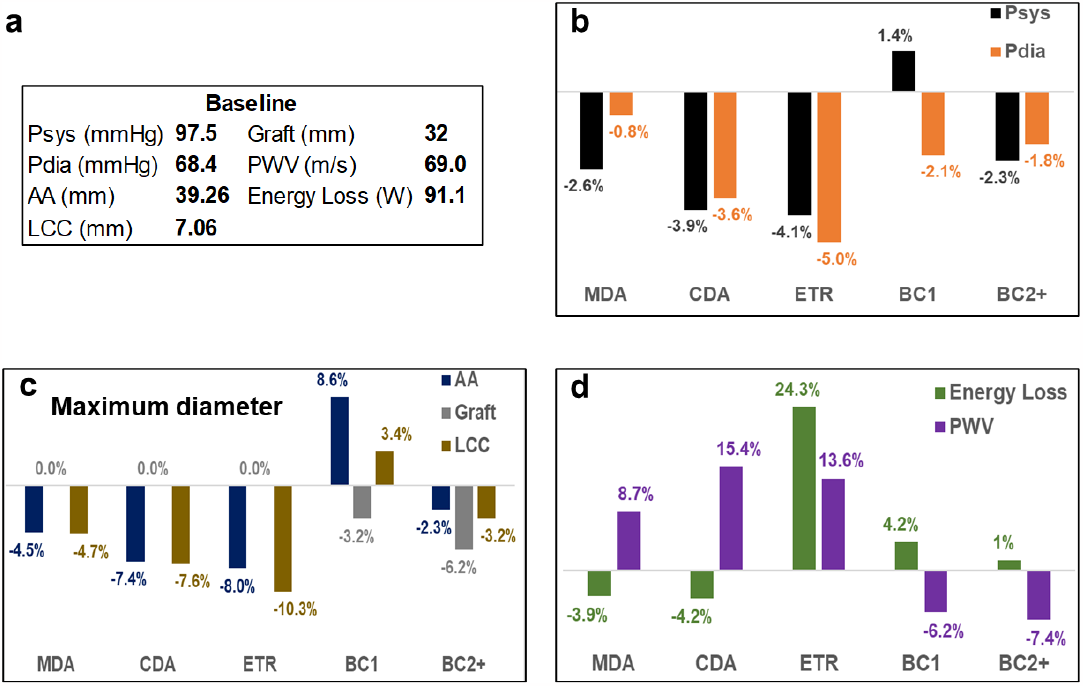
A Baseline case: inlet systolic and diastolic pressure, diameter at maximum displacement of the AA, left common carotid (LCC) and graft; PWV and EL. B, C and D Relative errors between the baseline and virtual intervention cases.

The pulse wave velocity (PWV) between the pressure peak at the inlet and the celiac trunk was calculated from the temporal difference in pressure wave peaks at a proximal and distal location in each case (Fig 3D). The PWV decreased by up to 15.4% in the cases where the graft was more rigid and increased up to 12 ms in BC2+, where the graft was the most compliant.

The maximum diameters of the AA, LCC and graft were compared between the baseline and the five additional virtual intervention cases (for clarity purposes, only the LCC is showed in Fig 3C as the displacements of the two other supra-aortic branches followed the same trend). The maximum diameters increased along with the pressures in the rigid graft cases, with maximum values found in the ETR case. The AA and supra-aortic branches maximum diameters were reduced in the BC1 case, while the maximum diameter of the compliant graft expanded by 3%. Maximum diameters at the three locations of interest of the BC2+ case were all larger than those to the baseline case, the compliant graft of BC2+ expanded by 6%.

The maximum increase in EL between the inlet and the outlets was observed in the CDA, while EL drastically decreased in the ETR case (Fig. 3D). EL was also slightly reduced in the BC1 case, while the change was negligible in BC2+.

### WSS-Based Indices

Contours of TAWSS and ECAP, capped between the critical ranges (0-5 Pa) and (0-1.4 Pa^-1^) according to literature [48][53] are plotted in Fig 4, 5 and 6. WSS distributions are similar among all cases, as the same inlet condition was applied and the geometries are similar. The PET, aneurysm and graft sutures are the main clinical regions of interest and hence differences in estimated indices between cases are illustrated therein. For compliteness, the BC1 and BC2+ cases are left on the different figures even if differences are negligeable.

**Fig 4.**
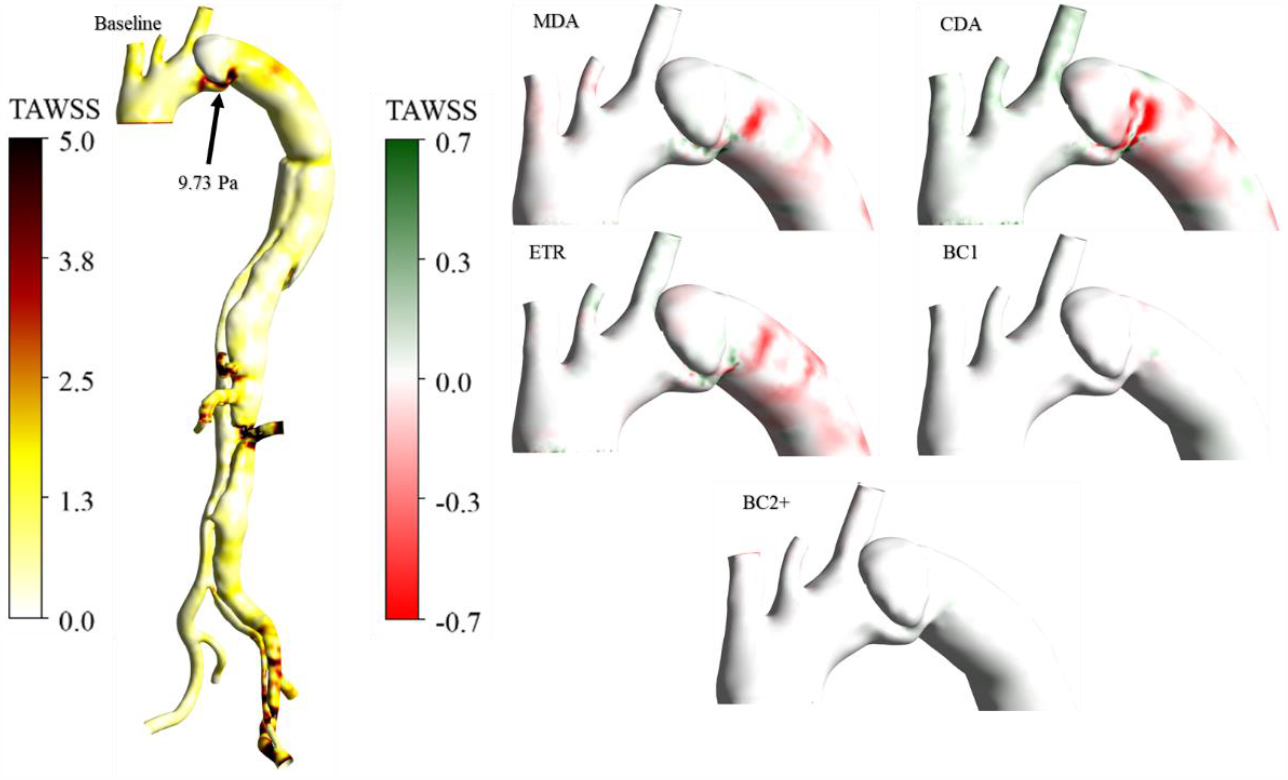
Front view of the TAWSS. On the left, values over 5 Pa are found at the PET, the abdominal arteries, and the left iliac of the baseline case. The black arrow indicates the maximum TAWSS at the PET. On the right, the TAWSS differences between the baseline and the five cases are shown. A zoom is made on the AA and aortic arch as regions of interest where the TAWSS is high.

**Fig 5.**
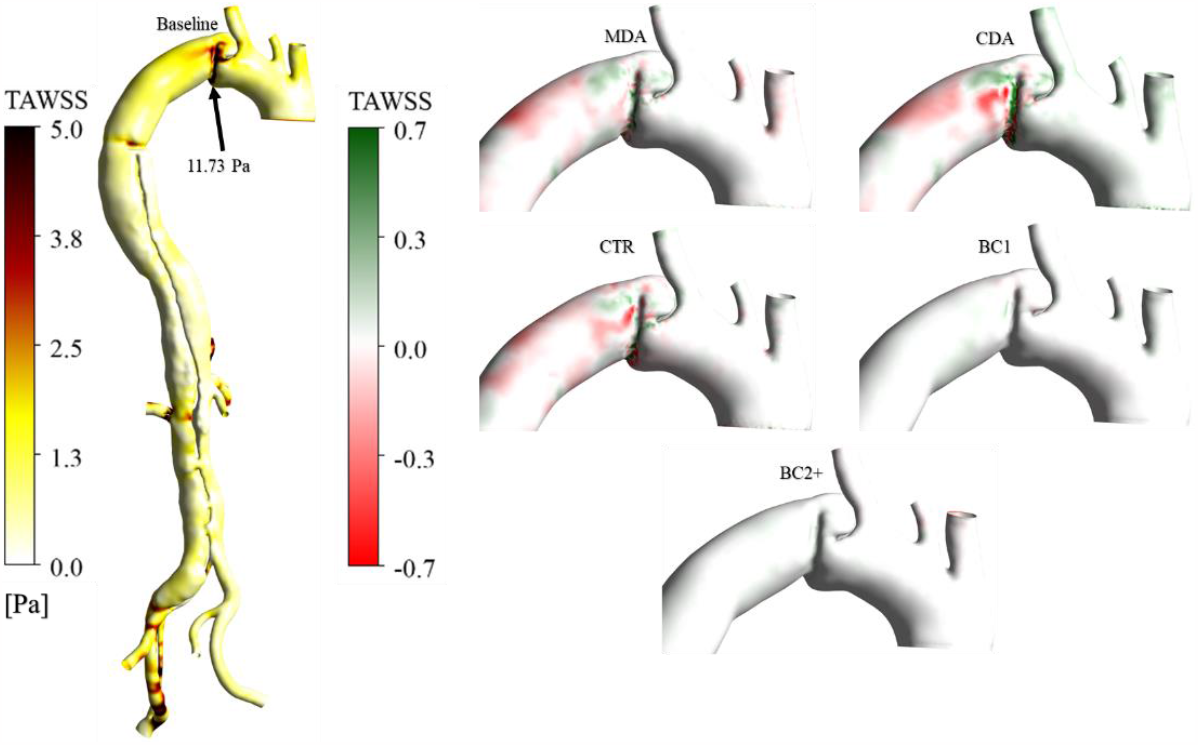
Front view of the TAWSS. On the left, values over 5 Pa are found at the PET and sutures with the graft of the baseline case. The black arrow indicates the maximum TAWSS at the PET. On the right, the TAWSS differences between the baseline and the five other cases are shown. A zoom is made on the AA and aortic arch as regions of interest where the TAWSS is high.

**Fig 6.**
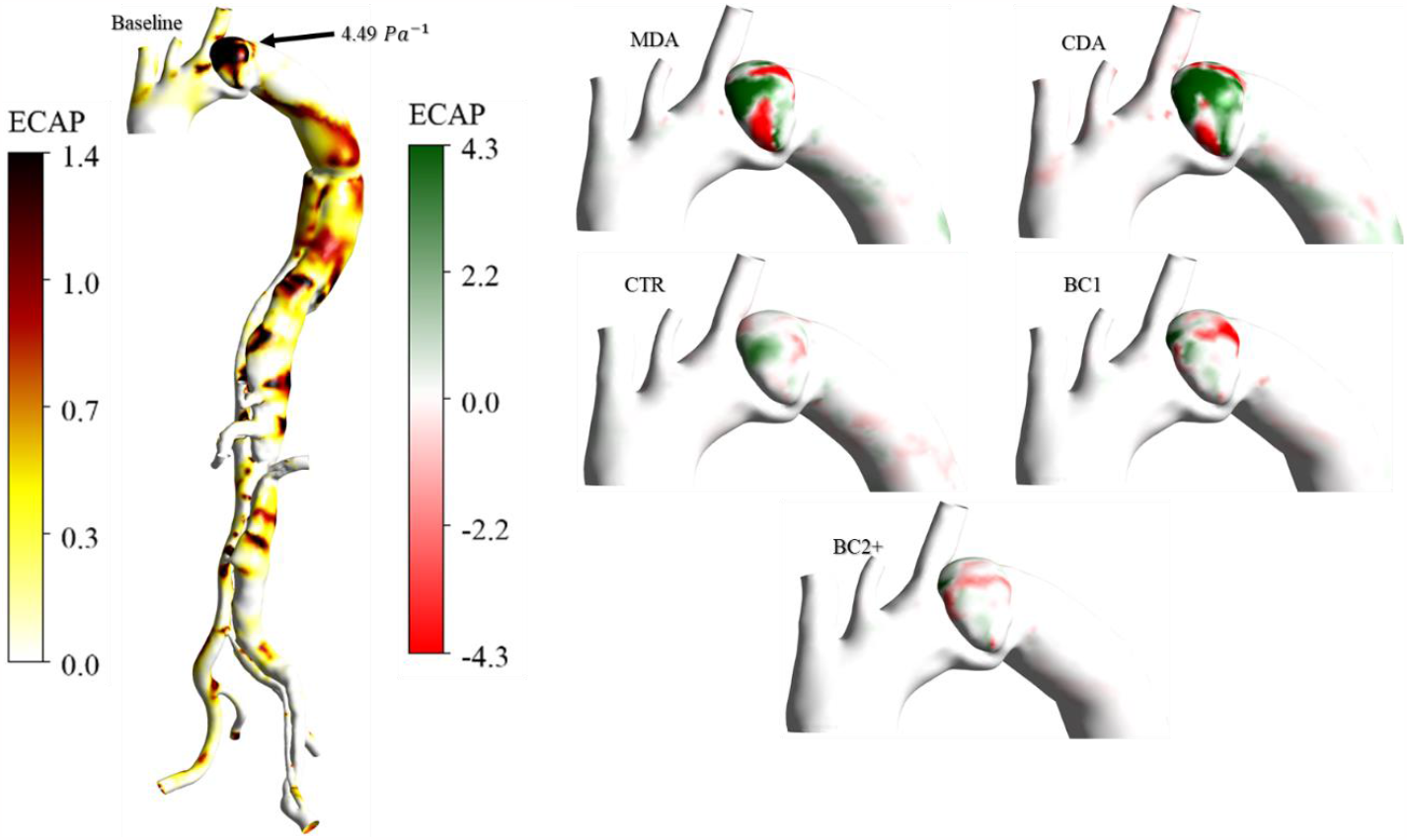
ECAP distributions, front view. On the left, ECAP absolute values for baseline case; values over 1.4 *Pa*^−*1*^ are noted in the aneurysm. The black arrow indicates the maximum ECAP value in the aneurysm. On the right, ECAP differences between the baseline and the five virtual cases.

High TAWSS is observed at the vinicity of PET, graft sutures, and abdominal and iliac arteries in every case due to high velocities in these locations, as observed in other studies [54][55][56]. Differences are primarily observed at the PET and graft sutures where TAWSS is high. The TAWSS maximum values at locations of interest are indicated on the baseline case in Fig 4 and Fig 5 by a black arrow. TAWSS marginally increases at the sutures and PET with a longer graft graft minimum and maximum differences are -0.8% and -1.37% at the PET and 1.87% and 2.28% at the sutures for the MDA and CDA cases, respectively. TAWSS distributions were qualitatively similar between the baseline and the ETR case. Normalised differences in BC1 and BC2+ were insignificant as these cases share identical boundary conditions and geometries.

ECAP values vary mostly within the aneurysm. They are generally over the critical value of 1.4 *Pa*^−1^ [57] with a maximum baseline value of 4.49 *Pa*^−1^, indicated by a black arrow in Fig 6. Critical ECAP values are also observed around the multiple re-entry tears proximal to the abdominal branches and the narrowing of the aortic lumens. Maximum differences in ECAP varied between 16% and 20% in the MDA and CDA cases, respectively. and were much smaller, within 7% in the ETR case. Similarly, to TAWSS distributions, ECAP distributions were very similar in BC1, BC2+ and baseline cases.

## Discussion

The blood pressure is controlled in TBAD patients, often using beta-blockers [58]. *P*_*sys,a*_ and *P*_*dia,a*_ are used as primary monitors of aortic and cardiovascular diseases; aortic pressure increase usually occurs after a TBAD [59]. Surgical Aortic repairs using grafts have been shown to lead to unfavourable, immediate and long-term cardiac outcomes, such as LV hypertrophy [60][61], due to increased aortic stifness and pressure.

### Pressure and Wall Displacement

In this study, aortic pressure increased with longer virtual grafts due to the associated increase in aortic stifness and impedance. Compared to the baseline simulation, *P*_*sys,a*_ and *P*_*dia,a*_ increased by approximately 4% in the MDA and CDA cases. In ETR, pressures remained close to those in MDA and CDA, while the pulse pressure decreased by 2.0% compared to the baseline case. The abdominal portion of the ETR case is grafted and has four circular outlets, which is considered ideal compared to the dissected and narrowed portion of the baseline, MDA and CDA cases. This most likely reduces the pressure waves reflection. Hence, even if the graft is longer and the aortic compliance smaller in the ETR case, the pressure did not increase as much as expected. In the BC1 case, the patient-specific compliant graft led to a decrease of 10.6% in pulse pressure. This suggests that a patient-specific compliant graft, may decrease the risk of LV hypertrophy. However, in BC2+, the pulse pressure increased by 3.6% compared to the baseline case. This suggests that adding too much compliance to the aorta can be detrimental and can increase aortic impedance. The implication is that the compliance mismatch between the graft and the aorta works in two ways; a graft more compliant than the natural aorta promotes as much a pressure increase as a rigid one.

The pressure wave transmission in an infinitely rigid straight tube is instantaneous. In cardiology, a stiffer aorta results in a higher PWV and is known to impair its ability to accomodate the cardiac output, acting to increase the LV load [62]. Due to the increased aortic rigidity and impedance introduced by longer grafts, which act to reduce volume compliance, we observed higher PWV. Similarly, compliant grafts that matched aortic compliance, reduced PWV between the inlet and the CT, thus reducing LV afterload. This difference in PWV reflects well the impact of the different grafting strategies; the more compliant the aorta, the larger the PWV due to the damping effect of the graft.

In recent rigid grafts studies, Rong et al. [63] and Nauta et al. [64] found increasing diameters in supra-aortic branches and DA after ascending and thoracic repairs. They noted an increase in pulse pressure and deformation of the AA and aortic arch, increasing the risk of dissection propagation or aneurysmal degeneration. Increased aortic distensibility is a risk factor for AD [65][66]. Our results show that the maximum displacements of the AA and supra-aortic branches increased with longer grafts. This phenomenon can be attributed to the formulation of the MBM, in which wall displacement is directly proportional to the local wall pressure difference and stiffness.Our findings indicate that longer grafts might increase the risk of dissection propagation and additional aortic tearing [66]. In the BC1 case, maximum displacements were reduced at the AA and supra-aortic branches due to the compliance of the graft, which also dilated and added aortic compliance, resulting in pressure reduction. The AA and aortic root axial displacement and motion are known risk factors of AD [67]. The buffer effect of the graft of the BC1 case may have potentially reduce the risk of ascending aortic aneurysm or additional tears in the arch or supra-aortic branches [68]. The BC2+ case exhibited higher displacements despite the graft been compliant. This is in line with the discussion as a more compliant graft than the adjacent tissue can increase the resistance to blood flow and introduce impedence mismatch, hence pressures and aortic wall displacements increase. This highlights that a too compliant graft can be as detrimental as a rigid one and that an optimum level of compliance exists to account for impact of compliance mismatch on hemodynamics.

### Energy Loss

EL is a marker of clinical interest used in cardiology and following visceral interventions, due to its association with poorer TBAD outcomes such as heart disease and heart failure [69][70]. EL has been used in CFD studies of virtual TEVAR [71] and idealised diseased aortas [72] using FSI. It was reported by Qiao et al. [71] that the interaction of the implanted graft and wall movement may be responsible for increased EL. In an FSI study comparing a pre-and post-TEVAR case of TBAD, van Bakel et al. [10] showed an increase in LV stroke work after the intervention. They concluded that the increaed aortic impedance and decreased aortic compliance between the endovascular stent and the aorta led to an increased LV afterload and proposed that compliant devices should be used. The same conclusions can be reached in terms of both stents and grafts as they are rigid devices that introduce a compliance mismatch with the aorta.

In our study, energy loss increased by approximately 4% in the MDA and CDA cases where grafted regions were more extensive. This suggests that the heart would need to exert more ejection force during each cardiac cycle. As mentioned in previous sections and discussed by Takeda et al. [73], increasing vascular stifness and aortic impedance increases the EL, thus increasing the risk of LV hypertrophy. However, the ETR case showed a remarkable reduction in EL of 24.3%. This suggests that replacing the total dissected portion in TBAD leads to a notable reduction in EL and may be a favourable surgical option if no other factors are considered. That been said, complete replacement of the aorta has been associated with serious negative consequences, such as spinal cord injury resulting in paraplegia, as most segmental arteries are no longer attached to the aorta [74]. Also, in the case of a more extensive dissection, kidneys adjust to a physiologically abnormal level of perfusion. Therefore, recovering a physiologically typical flow split after surgery may lead to the deterioration of renal function [75]. Additionally, due to the added aortic compliance introduced by the use of a compliant graft, EL decreased by 4% in the BC1 case compared to the baseline. This phenomenon should reduce the risk of LV hypertrophy [73]. Counterintuitively the BC2+ case resulted in a slight elevation in EL was simulated in the BC2+ case compared to the baseline. This emphasises that an increase in aortic compliance can lead to an increase of aortic impedance leading to an increase in pressure and EL. These findings support that a graft with excessive compliance is also unfavourable for the treatment of TBAD [76].

### WSS-Based Indices

High TAWSS values are commonly found in narrowed regions such as the PET and re-entry tears due to higher velocity gradients in these regions [40]. Our results showed qualitatively similar TAWSS distributions across all cases. The major normalised differences were found between the baseline and MDA and CDA cases near the PET and graft sutures. High TAWSS has been linked with aortic wall degeneration and rupture [77]. However, it was reported by Alimohammadi et al. [31], that in regions of high TAWSS, minimal differences are observed between equivalent rigid and compliant simulations. Similarly, our results show that normalised relative differences between the baseline case, BC1 and BC2+, were negligible.

Higher TAWSS was observed in the MDA and CDA at PET compared to the baseline case; this could indicate a higher risk of aortic growth, dissection and tear expansion [78][79][80]. TAWSS was slightly lower at the sutures, which may suggest that the risk of rupture is also lower. However, values were still above the critical value of 5 Pa, which has been linked to dissection tear initiation [53]. The ETR case exhibited a TAWSS distribution nearly identical to the baseline case. This resulted in the same findings regarding potential risk areas, such as the PET, sutures, and aneurysms.

The differences in TAWSS and ECAP between the baseline case and BC1 and BC2+ were minimal across the whole geometry. This indicates that using a compliant graft does not significantly impact the distribution of WSS compared to a rigid graft when applying the same boundary conditions to the same geometry.

ECAP was high throughout the aneurysmal region in the baseline case. High ECAP may indicate regions with an elevated risk of atherosclerotic plaque formation and calcification, a known risk factor for aortic rupture commonly found in TBAD [81]. Noticeable differences in ECAP were observed between the baseline and the CDA and MDA cases, with mximum increases of 16% and 20% at the aneurysm, respectively. This indicates that graft length directly affects ECAP and thus may influence aortic wall remodelling and disease progression.

### Compliant Biomimicking Grafts

Research has demonstrated significant progress in 3D bioprinting technology. In recent years, tissue analogues for aortic valves or blood vessels, have been successfully produced [82]. However, biomimicking technologies for compliant tissues have been mostly applied to smaller vessels [83]. Reproducing or mimicking the characteristics of the aorta remains complex and costly and has been scarcely reported [84]. Our findings suggest that compliant grafts may benefit TBAD patients after OS by reducing EL and thus reducing the risk of LV hypertrophy and heart failure. Combining *in silico* virtual grafting and *in vivo* imaging data, 3D bioprinting technology may facilitate further research and attract graft and stent manufacturers interest in this direction.

### Limitations

In this study, we investigated the impact of graft length and compliance in a patient-specific case of chronic TBAD using routinely acquired clinical data includinglimited pre-operative 2DMRI and Cine-MRI data. Using pre-operative data may introduce inaccuracies in post-intervention virtual scenarios due to changes in inlet flow rate and aortic wall compliance after the intervention. However, previous research by Pirola et al. [85] demonstrated the feasibility of using preoperative data to tune postoperative boundary conditions by using post-intervention invasive aortic pressure measurements acquired during a follow-up, and showed overall acceptable agreement with their simulated post-intervention pressure. Our results suggest that this methodology can be valuable in the absence of clinical data during the follow-up of TBAD patients with graft.

Future work will endeavour to incorporate 4DMR and Cine-MRI datasets to improve the accuracy of the simulations. These datasets will enable a more comprehensive description of the haemodynamics and wall movement of the aorta and facilitate further validation of the simulations. This approach may lead to well validated simulations informed by rich datasets, with the potential to capture more accurately key haemodynamic variables of interest.

## Conclusions

This study simulated a patient-specific post-operative case of TBAD and explored the impact of different surgical strategies via virtual grafting. Specifically, we conducted five simulations, i.e., virtual interventions, including three virtual surgeries using varying graft sizes and two cases with compliant graft. To the authors knowledge, this study is the first investigation in the literature aiming to evaluate the influence of various potential surgical strategies for TBAD on key haemodynamic markers, WSS distribution, and LV workload, considering the effects of graft length and compliance.

Our findings suggest that reducing aortic volume compliance by increasing the length of rigid grafts increases pressure and EL. Furthermore, a graft with a compliance matching the natural aortic compliance of the patient lowered the inlet and pulse pressures and EL. ECAP differed greatly between different grafting strategies within the aneurysmal region of the dissection, with maximum increases of 16% and 20% between the baseline case and MDA and CDA, respectively. In conclusion, optimal graft selection cannot be determined without considering the morphology and condition of the aorta of each individual patient. Exploring various virtual grafting strategies via patient-specific simulations can aid this process as illustrated in this study. graft manufacturers should consider developing biomimetic grafts to reduce the risk of LV hypertrophy and heart failure in future TBAD patients.

## Supporting information

Manuscript

## Data Availability

All data produced in the present study are available upon reasonable request to the authors

## Abbreviations

AD: Aortic Dissection
BC1: Baseline compliant one
BC2+: Baseline compliant two plus
BT: Braciocephalic trunk
CDA: Complete descending aorta
CT: Celiac trunk
CTA: Computed tomography angiography
DA: Descending aorta
ETR: Entire thoracoabdominal replacement
FSI: Fluid-structure interaction
LCC: Left common carotid
LEI: Left exterior iliac
LII: Left interior iliac
LR: Left renal
LSA: Left subclavian
LV: Left ventricular
MDA: Mid-descending aorta
MBM: Moving boundary method
OS: Open surgery
PET: Primary entry tear
PWV: Pulse wave velocity
REI: Right exterior iliac
RII: Right interior iliac
RR: Right renal
SMA: Superior mesenteric
TEVAR: Thoracic endovascular aortic repair
WSS: Wall shear stress
2DMRI: 2D-flow magnetic resonance imaging
4DMRI: 4D-flow magnetic resonance imaging

## Acknowledgements

The authors would like to thank the Department of Computer Science of University College London for the high-performance computing cluster resources used to perform the simulations.

## References

[1] J. M. Trahanas, O. A. Jarral, C. Long, and G. C. Hughes, “Management of chronic type B aortic dissection,” Vessel Plus, vol. 6, 2022, doi: 10.20517/2574-1209.2021.125.

[2] M. L. Williams et al., “Thoracic endovascular repair of chronic type B aortic dissection: A systematic review,” Ann Cardiothorac Surg, vol. 11, no. 1, pp. 1–15, 2022, doi: 10.21037/ACS-2021-TAES-25.

[3] R. W. Hsieh et al., “Comparison of type B dissection by open, endovascular, and medical treatments,” J Vasc Surg, vol. 70, no. 6, pp. 1792–1800.e3, Dec. 2019, doi: 10.1016/j.jvs.2019.02.062.

[4] P. P. Goodney et al., “Cardiovascular Surgery Survival After Open Versus Endovascular Thoracic Aortic Aneurysm Repair in an Observational Study of the Medicare Population,” 2011, doi: 10.1161/CIRCULATIONAHA.111.033944/-/DC1.

[5] D. H. Tian, R. P. De Silva, T. Wang, T. D. Yan, and P. of Cardiovascular, “Open surgical repair for chronic type B aortic dissection: a systematic review Systematic Review Background,” Ann Cardiothorac Surg, vol. 3, no. 4, pp. 340–350, 2014, doi: 10.3978/j.issn.2225-319X.2014.07.10.

[6] L. di Tommaso, R. Giordano, E. di Tommaso, and G. Iannelli, “Endovascular treatment for chronic type B aortic dissection: Current opinions,” Journal of Thoracic Disease, vol. 10. AME Publishing Company, S978–S982, Apr. 01, 2018. doi: 10.21037/jtd.2018.03.145.

[7] S. A. Son, H. Jung, and J. Y. Cho, “Long-term outcomes of intervention between open repair and endovascular aortic repair for descending aortic pathologies: a propensity-matched analysis,” BMC Surg, vol. 20, no. 1, pp. 1–13, 2020, doi: 10.1186/s12893-020-00923-4.

[8] C. Spadaccio et al., “Old Myths, New Concerns: the Long-Term Effects of Ascending Aorta Replacement with Dacron Grafts. Not All That Glitters Is Gold,” J Cardiovasc Transl Res, vol. 9, no. 4, pp. 334–342, 2016, doi: 10.1007/s12265-016-9699-8.

[9] S. Y. Kim, T. J. Hinkamp, W. R. Jacobs, R. C. Lichtenberg, H. Posniak, and R. Pifarr6, “Effect of an Inelastic Aortic Synthetic Vascular Graft on Exercise Hemodynamics,” 1995.

[10] T. M. J. Van Bakel et al., “Cardiac remodelling following thoracic endovascular aortic repair for descending aortic aneurysms,” European Journal of Cardio-thoracic Surgery, vol. 55, no. 6, pp. 1061–1070, 2019, doi: 10.1093/ejcts/ezy399.

[11] N. Brown, “IMPEDANCE MATCHING AT ARTERIAL BIFURCATIONS,” 1993.

[12] G. M. London and B. Pannier, “Arterial functions: How to interpret the complex physiology,” Nephrology Dialysis Transplantation, vol. 25, no. 12. xpp. 3815–3823, Dec. 2010. doi: 10.1093/ndt/gfq614.

[13] G. F. Mitchell et al., “Arterial stiffness, pressure and flow pulsatility and brain structure and function: The Age, Gene/Environment Susceptibility-Reykjavik Study,” Brain, vol. 134, no. 11, pp. 3398–3407, 2011, doi: 10.1093/brain/awr253.

[14] J. Chung et al., “Energy loss, a novel biomechanical parameter, correlates with aortic aneurysm size and histopathologic findings,” in Journal of Thoracic and Cardiovascular Surgery, Mosby Inc., 2014, 1082–1089. doi: 10.1016/j.jtcvs.2014.06.021.

[15] C. Tsioufis et al., “Left ventricular diastolic dysfunction is accompanied by increased aortic stiffness in the early stages of essential hypertension: A TDI approach,” J Hypertens, vol. 23, no. 9, pp. 1745–1750, 2005, doi: 10.1097/01.hjh.0000174394.57644.69.

[16] C. V. Ioannou et al., “Left ventricular hypertrophy induced by reduced aortic compliance,” J Vasc Res, vol. 46, no. 5, pp. 417–425, Aug. 2009, doi: 10.1159/000194272.

[17] W. W. Nichols, S. J. Denardo, I. B. Wilkinson, C. M. McEniery, J. Cockcroft, and M. F. O’Rourke, “Effects of arterial stiffness, pulse wave velocity, and wave reflections on the central aortic pressure waveform,” J Clin Hypertens, vol. 10, no. 4, 295–303, 2008, doi: 10.1111/j.1751-7176.2008.04746.x.

[18] C. A. Valencia-Hernández et al., “Aortic Pulse Wave Velocity as Adjunct Risk Marker for Assessing Cardiovascular Disease Risk: Prospective Study,” Hypertension, vol. 79, no. 4, pp. 836–843, Apr. 2022, doi: 10.1161/HYPERTENSIONAHA.121.17589.

[19] E. M. Isselbacher et al., “2022 ACC/AHA Guideline for the Diagnosis and Management of Aortic Disease: A Report of the American Heart Association/American College of Cardiology Joint Committee on Clinical Practice Guidelines,” Circulation, vol. 146, no. 24. Lippincott Williams and Wilkins, pp. E334–E482, Dec. 13, 2022. doi: 10.1161/CIR.0000000000001106.

[20] A. Evangelista et al., “Multimodality imaging in thoracic aortic diseases: a clinical consensus statement from the European Association of Cardiovascular Imaging and the European Society of Cardiology working group on aorta and peripheral vascular diseases,” Eur Heart J Cardiovasc Imaging, Mar. 2023, doi: 10.1093/ehjci/jead024.

[21] F. Rengier et al., “Impact of an Aortic Nitinol Stent Graft on Flow Measurements by Time-resolved Three-dimensional Velocity-encoded MRI,” Acad Radiol, vol. 19, no. 3, 274–280, 2012, doi: 10.1016/j.acra.2011.10.025.

[22] C. W. Chen et al., “Aortic dissection assessment by 4D phase-contrast MRI with hemodynamic parameters: The impact of stent type,” Quant Imaging Med Surg, vol. 11, no. 2, pp. 490–501, Feb. 2021, doi: 10.21037/QIMS-20-670.

[23] M. Markl, “How well does an automated approach calculate and visualize blood flow vorticity at 4d flow mri?,” Radiology: Cardiothoracic Imaging, vol. 2, no. 1. Radiological Society of North America Inc., Feb. 01, 2020. doi: 10.1148/ryct.2020190233.

[24] M. Markl et al., “Advanced flow MRI: emerging techniques and applications,” Clinical Radiology, vol. 71, no. 8. W.B. Saunders Ltd, pp. 779–795, Aug. 01, 2016. doi: 10.1016/j.crad.2016.01.011.

[25] C. W. Ong et al., “Computational Fluid Dynamics Modeling of Hemodynamic Parameters in the Human Diseased Aorta: A Systematic Review,” Annals of Vascular Surgery, vol. 63. Elsevier Inc., pp. 336–381, Feb. 01, 2020. doi: 10.1016/j.avsg.2019.04.032.

[26] K. Lu et al., “Computational Study of Fenestration and Parallel Grafts Used in TEVAR of Aortic Arch Aneurysms,” pp. 0–3, doi: 10.1002/cnm.3664.

[27] K. Wang, D. Li, D. Yuan, J. Zhao, T. Zheng, and Y. Fan, “A computational fluid study on hemodynamics in visceral arteries in a complicated type B aortic dissection after thoracic endovascular repair,” Med Nov Technol Devices, vol. 9, no. December 2020, p. 100054, 2021, doi: 10.1016/j.medntd.2020.100054.

[28] N. Westerhof, J. W. Lankhaar, and B. E. Westerhof, “The arterial windkessel,” Medical and Biological Engineering and Computing, vol. 47, no. 2. pp. 131–141, 2009. doi: 10.1007/s11517-008-0359-2.

[29] C. H. Armour et al., “The influence of inlet velocity profile on predicted flow in type B aortic dissection,” Biomech Model Mechanobiol, vol. 20, no. 2, pp. 481–490, 2021, doi: 10.1007/s10237-020-01395-4.

[30] A. Boccadifuoco, A. Mariotti, S. Celi, N. Martini, and M. V. Salvetti, “Impact of uncertainties in outflow boundary conditions on the predictions of hemodynamic simulations of ascending thoracic aortic aneurysms,” Comput Fluids, vol. 165, pp. 96–115, Mar. 2018, doi: 10.1016/j.compfluid.2018.01.012.

[31] M. Alimohammadi, J. M. Sherwood, M. Karimpour, O. Agu, S. Balabani, and V. Díaz-Zuccarini, “Aortic dissection simulation models for clinical support: Fluid-structure interaction vs. rigid wall models,” Biomed Eng Online, vol. 14, no. 1, Apr. 2015, doi: 10.1186/s12938-015-0032-6.

[32] G. Lee, Y. Lee, and T. Kim, “Fluid-structure interaction simulation of visceral perfusion and impact of different cannulation methods on aortic dissection,” pp. 1–23.

[33] M. Bonfanti, S. Balabani, M. Alimohammadi, O. Agu, S. Homer-Vanniasinkam, and V. Díaz-Zuccarini, “A simplified method to account for wall motion in patient-specific blood flow simulations of aortic dissection: Comparison with fluid-structure interaction,” Med Eng Phys, vol. 58, pp. 72–79, Aug. 2018, doi: 10.1016/j.medengphy.2018.04.014.

[34] K. Bäumler et al., “Fluid–structure interaction simulations of patient-specific aortic dissection,” Biomech Model Mechanobiol, vol. 19, no. 5, pp. 1607–1628, 2020, doi: 10.1007/s10237-020-01294-8.

[35] G. Nannini et al., “Aortic hemodynamics assessment prior and after valve sparing reconstruction: A patient-specific 4D flow-based FSI model,” Comput Biol Med, vol. 135, no. April, p. 104581, 2021, doi: 10.1016/j.compbiomed.2021.104581.

[36] A. Aghilinejad, H. Wei, G. A. Magee, and N. M. Pahlevan, “Model-Based Fluid-Structure Interaction Approach for Evaluation of Thoracic Endovascular Aortic Repair Endograft Length in Type B Aortic Dissection,” vol. 10, no. June, pp. 1–14, 2022, doi: 10.3389/fbioe.2022.825015.

[37] B. A. Craven, E. G. Paterson, G. S. Settles, and M. J. Lawson, “Development and verification of a high-fidelity computational fluid dynamics model of canine nasal airflow,” J Biomech Eng, vol. 131, no. 9, pp. 1–11, 2009, doi: 10.1115/1.3148202.

[38] Z. Cheng, N. B. Wood, R. G. J. Gibbs, and X. Y. Xu, “Geometric and Flow Features of Type B Aortic Dissection: Initial Findings and Comparison of Medically Treated and Stented Cases,” Ann Biomed Eng, vol. 43, no. 1, pp. 177–189, 2015, doi: 10.1007/s10439-014-1075-8.

[39] M. Amanuma, R. H. Mohiaddin, M. Hasegawa, A. Heshiki, and D. B. Longmore, “Abdominal aorta: characterisation of blood flow and measurement of its regional distribution by cine magnetic resonance phase-shift velocity mapping,” Eur Radiol, vol. 2, no. 6, pp. 559–564, 1992, doi: 10.1007/BF00187552.

[40] M. Bonfanti, G. Franzetti, G. Maritati, S. Homer-Vanniasinkam, S. Balabani, and V. Díaz-Zuccarini, “Patient-specific haemodynamic simulations of complex aortic dissections informed by commonly available clinical datasets,” Med Eng Phys, vol. 71, pp. 45–55, Sep. 2019, doi: 10.1016/j.medengphy.2019.06.012.

[41] C. Stokes et al., “A novel MRI-based data fusion methodology for efficient, personalised, compliant simulations of aortic haemodynamics,” J Biomech, vol. 129, no. September, p. 110793, 2021, doi: 10.1016/j.jbiomech.2021.110793.

[42] G. Tomaiuolo, A. Carciati, S. Caserta, and S. Guido, “Blood linear viscoelasticity by small amplitude oscillatory flow,” Rheol Acta, vol. 55, no. 6, pp. 485–495, 2016, doi: 10.1007/s00397-015-0894-3.

[43] J. Peacock, T. Jones, C. Tock, and R. Lutz, “The onset of turbulence in physiological pulsatile flow in a straight tube,” Exp Fluids, vol. 24, no. 1, pp. 1–9, 1998, doi: 10.1007/s003480050144.

[44] N. Cagney and S. Balabani, “Influence of Shear-Thinning Rheology on the Mixing Dynamics in Taylor-Couette Flow,” Chem Eng Technol, vol. 42, no. 8, pp. 1680–1690, 2019, doi: 10.1002/ceat.201900015.

[45] J. Chung et al., “Energy loss, a novel biomechanical parameter, correlates with aortic aneurysm size and histopathologic findings,” Journal of Thoracic and Cardiovascular Surgery, vol. 148, no. 3, pp. 1082–1089, 2014, doi: 10.1016/j.jtcvs.2014.06.021.

[46] A. R. Babu, A. G. Byju, and N. Gundiah, “Biomechanical Properties of Human Ascending Thoracic Aortic Dissections,” J Biomech Eng, vol. 137, no. 8, Aug. 2015, doi: 10.1115/1.4030752.

[47] Y. Qiao, K. Luo, and J. Fan, “Component quantification of aortic blood flow energy loss using computational fluid-structure interaction hemodynamics,” Comput Methods Programs Biomed, vol. 221, p. 106826, 2022, doi: 10.1016/j.cmpb.2022.106826.

[48] Z. Sun and T. Chaichana, “A systematic review of computational fluid dynamics in type B aortic dissection,” Int J Cardiol, vol. 210, pp. 28–31, 2016, doi: 10.1016/j.ijcard.2016.02.099.

[49] D. Gallo, G. De Santis, D. Tresoldi, I. National, and R. Ponzini, “On the Use of In Vivo Measured Flow Rates as Boundary Conditions for Image-Based Hemodynamic Models of the Human Aorta : Implications for Indicators of Abnormal Flow On the Use of In Vivo Measured Flow Rates as Boundary Conditions for Image-Based Hemodyn,” no. May, 2014, doi: 10.1007/s10439-011-0431-1.

[50] P. Di Achille, G. Tellides, C. A. Figueroa, and J. D. Humphrey, “A haemodynamic predictor of intraluminal thrombus formation in abdominal aortic aneurysms,” Proceedings of the Royal Society A: Mathematical, Physical and Engineering Sciences, vol. 470, no. 2172, 2014, doi: 10.1098/rspa.2014.0163.

[51] L. J. Kelsey, J. T. Powell, P. E. Norman, K. Miller, and B. J. Doyle, “A comparison of hemodynamic metrics and intraluminal thrombus burden in a common iliac artery aneurysm,” Int J Numer Method Biomed Eng, vol. 33, no. 5, May 2017, doi: 10.1002/cnm.2821.

[52] F. Kohno, T. Kumada, M. Kambayashi, W. Hayashida, N. Ishikawa, and S. Sasayama, “Change in aortic end-systolic pressure by alterations in loading sequence and its relation to left ventricular isovolumic relaxation,” Circulation, vol. 93, no. 11, pp. 2080–2087, 1996, doi: 10.1161/01.CIR.93.11.2080.

[53] L. Peng et al., “Patient-specific Computational Hemodynamic Analysis for Interrupted Aortic Arch in an Adult: Implications for Aortic Dissection Initiation,” Sci Rep, vol. 9, no. 1, Dec. 2019, doi: 10.1038/s41598-019-45097-z.

[54] D. Chen, M. Müller-Eschner, D. Kotelis, D. Böckler, Y. Ventikos, and H. Von Tengg-Kobligk, “A longitudinal study of Type-B aortic dissection and endovascular repair scenarios: Computational analyses,” Med Eng Phys, vol. 35, no. 9, pp. 1321–1330, 2013, doi: 10.1016/j.medengphy.2013.02.006.

[55] I. Wee, C. W. Ong, N. Syn, and A. Choong, “Computational Fluid Dynamics and Aortic Dissections: Panacea or Panic?,” Vascular and Endovascular Review, vol. 1, no. 1, pp. 27–29, 2018, doi: 10.15420/ver.2018.8.2.

[56] C. H. Armour, C. Menichini, K. Milinis, R. G. J. Gibbs, and X. Y. Xu, “Location of Reentry Tears Affects False Lumen Thrombosis in Aortic Dissection Following TEVAR,” Journal of Endovascular Therapy, vol. 27, no. 3, pp. 396–404, Jun. 2020, doi: 10.1177/1526602820917962.

[57] A. Deyranlou, C. A. Miller, A. Revell, and A. Keshmiri, “Effects of Ageing on Aortic Circulation During Atrial Fibrillation; a Numerical Study on Different Aortic Morphologies,” Ann Biomed Eng, 2021, doi: 10.1007/s10439-021-02744-9.

[58] T. Suzuki, K. A. Eagle, E. Bossone, A. Ballotta, J. B. Froehlich, and E. M. Isselbacher, “Medical management in type B aortic dissection.,” Ann Cardiothorac Surg, vol. 3, no. 4, pp. 413–7, 2014, doi: 10.3978/j.issn.2225-319X.2014.07.01.

[59] J. C. Zhou et al., “Intensive blood pressure control in patients with acute type B aortic dissection (RAID): Study protocol for randomized controlled trial,” J Thorac Dis, vol. 9, no. 5, pp. 1369–1374, 2017, doi: 10.21037/jtd.2017.03.180.

[60] Y. Ikeno, V. T. T. Truong, A. Tanaka, and S. K. Prakash, “The Effect of Ascending Aortic Repair on Left Ventricular Remodeling,” American Journal of Cardiology, vol. 182, no. Lv, pp. 89–94, 2022, doi: 10.1016/j.amjcard.2022.07.027.

[61] S. Sultan, Y. Acharya, O. Soliman, J. C. Parodi, and N. Hynes, “TEVAR and EVAR, the unknown knowns of the cardiovascular hemodynamics; and the immediate and long-term consequences of fabric material on major adverse clinical outcome,” Front Surg, vol. 9, no. August, pp. 1–8, 2022, doi: 10.3389/fsurg.2022.940304.

[62] M. F. O’ROURKE, J. V. Blazek, C. L. Morreels, and L. J. Krovetz, “Pressure Wave Transmission along the Human Aorta,” Circ Res, vol. 23, no. 4, pp. 567–579, 1968, doi: 10.1161/01.res.23.4.567.

[63] L. Q. Rong et al., “Immediate Impact of Prosthetic Graft Replacement of the Ascending Aorta on Circumferential Strain in the Descending Aorta,” European Journal of Vascular and Endovascular Surgery, vol. 58, no. 4, pp. 521–528, 2019, doi: 10.1016/j.ejvs.2019.05.003.

[64] F. J. H. Nauta et al., “Impact of thoracic endovascular aortic repair on pulsatile circumferential and longitudinal strain in patients with aneurysm,” Journal of Endovascular Therapy, vol. 24, no. 2, pp. 281–289, 2017, doi: 10.1177/1526602816687086.

[65] H. W. L. de Beaufort et al., “Extensibility and Distensibility of the Thoracic Aorta in Patients with Aneurysm,” European Journal of Vascular and Endovascular Surgery, vol. 53, no. 2, pp. 199–205, 2017, doi: 10.1016/j.ejvs.2016.11.018.

[66] G. H. W. van Bogerijen et al., “Importance of dynamic aortic evaluation in planning TEVAR.,” Ann Cardiothorac Surg, vol. 3, no. 3, pp. 300–6, 2014, doi: 10.3978/j.issn.2225-319X.2014.04.05.

[67] C. J. Beller, M. R. Labrosse, M. J. Thubrikar, and F. Robicsek, “Role of Aortic Root Motion in the Pathogenesis of Aortic Dissection,” Circulation, vol. 109, no. 6, pp. 763–769, Feb. 2004, doi: 10.1161/01.CIR.0000112569.27151.F7.

[68] K. A. Wilson, A. J. Lee, P. R. Hoskins, F. G. R. Fowkes, C. V. Ruckley, and A. W. Bradbury, “The relationship between aortic wall distensibility and rupture of infrarenal abdominal aortic aneurysm,” J Vasc Surg, vol. 37, no. 1, pp. 112–117, Jan. 2003, doi: 10.1067/mva.2003.40.

[69] F. M. Rijnberg et al., “Energetics of blood flow in cardiovascular disease: Concept and clinical implications of adverse energetics in patients with a fontan circulation,” Circulation, vol. 137, no. 22, pp. 2393–2407, 2018, doi: 10.1161/CIRCULATIONAHA.117.033359.

[70] M. Jozwiak et al., “Improved estimation of cardiac power output by including pulsatile power,” British Journal of Anaesthesia, vol. 125, no. 3. Elsevier Ltd, pp. e267–e269, Sep. 01, 2020. doi: 10.1016/j.bja.2020.05.010.

[71] Y. Qiao, L. Mao, Y. Ding, T. Zhu, K. Luo, and J. Fan, “Fluid-structure interaction: Insights into biomechanical implications of endograft after thoracic endovascular aortic repair,” Comput Biol Med, vol. 138, no. September, p. 104882, 2021, doi: 10.1016/j.compbiomed.2021.104882.

[72] Y. Qiao, L. Mao, Y. Ding, T. Zhu, K. Luo, and J. Fan, “Fluid-structure interaction: Insights into biomechanical implications of endograft after thoracic endovascular aortic repair,” Comput Biol Med, vol. 138, no. July, p. 104882, 2021, doi: 10.1016/j.compbiomed.2021.104882.

[73] Y. Takeda et al., “Endovascular aortic repair increases vascular stiffness and alters cardiac structure and function,” Circulation Journal, vol. 78, no. 2, pp. 322–328, 2014, doi: 10.1253/circj.CJ-13-0877.

[74] D. Petroff et al., “Paraplegia prevention in aortic aneurysm repair by thoracoabdominal staging with ‘minimally invasive staged segmental artery coil embolisation’ (MIS2ACE): Trial protocol for a randomised controlled multicentre trial,” BMJ Open, vol. 9, no. 3, 2019, doi: 10.1136/bmjopen-2018-025488.

[75] T. Urbanek, G. Biolik, W. Zelawski, B. Hapeta, M. Jusko, and W. Kuczmik, “The risk of renal function deterioration in abdominal aortic stent graft patients with and without previous kidney function failure – an analysis of risk factors,” Pol J Radiol, vol. 85, no. 1, pp. e643–e649, 2020, doi: 10.5114/PJR.2020.102194.

[76] M. Tang, D. Eliathamby, M. Ouzounian, C. A. Simmons, and J. C. Y. Chung, “Dependency of energy loss on strain rate, strain magnitude and preload: Towards development of a novel biomarker for aortic aneurysm dissection risk,” J Mech Behav Biomed Mater, vol. 124, Dec. 2021, doi: 10.1016/j.jmbbm.2021.104736.

[77] M. Y. Salmasi et al., “High Wall Shear Stress can Predict Wall Degradation in Ascending Aortic Aneurysms: An Integrated Biomechanics Study,” Front Bioeng Biotechnol, vol. 9, no. October, pp. 1–13, 2021, doi: 10.3389/fbioe.2021.750656.

[78] E. K. Shang et al., “Use of computational fluid dynamics studies in predicting aneurysmal degeneration of acute type B aortic dissections,” in Journal of Vascular Surgery, Mosby Inc., Aug. 2015, pp. 279–284. doi: 10.1016/j.jvs.2015.02.048.

[79] Y. Zhu et al., “Association of hemodynamic factors and progressive aortic dilatation following type A aortic dissection surgical repair,” Sci Rep, vol. 11, no. 1, Dec. 2021, doi: 10.1038/s41598-021-91079-5.

[80] O. Mutlu, H. E. Salman, H. Al-Thani, A. El-Menyar, U. A. Qidwai, and H. C. Yalcin, “How does hemodynamics affect rupture tissue mechanics in abdominal aortic aneurysm: Focus on wall shear stress derived parameters, time-averaged wall shear stress, oscillatory shear index, endothelial cell activation potential, and relative residence time,” Comput Biol Med, p. 106609, Mar. 2023, doi: 10.1016/j.compbiomed.2023.106609.

[81] P. N. Jonathan Golledge, “Vascular grafts.,” Expert Rev Cardiovasc Ther, vol. 1, no. 4, 81–594, 2003, doi: 10.1586/14779072.1.4.581.

[82] A. Khanna, B. P. Oropeza, and N. F. Huang, “Engineering Spatiotemporal Control in Vascularized Tissues,” Bioengineering, vol. 9, no. 10. MDPI, Oct. 01, 2022. doi: 10.3390/bioengineering9100555.

[83] M. J. Moreno, A. Ajji, D. Mohebbi-Kalhori, M. Rukhlova, A. Hadjizadeh, and M. N. Bureau, “Development of a compliant and cytocompatible micro-fibrous polyethylene terephthalate vascular scaffold,” J Biomed Mater Res B Appl Biomater, vol. 97 B, no. 2, pp. 201–214, 2011, doi: 10.1002/jbm.b.31774.

[84] J. Chlupác, E. Filová, and L. Bačáková, “Blood vessel replacement: 50 years of development and tissue engineering paradigms in vascular surgery,” Physiol Res, vol. 58, no. SUPPL.2, pp. 119–140, 2009, doi: 10.33549/physiolres.931918.

[85] S. Pirola et al., “4-D Flow mri-based computational analysis of blood flow in patient-specific aortic dissection,” IEEE Trans Biomed Eng, vol. 66, no. 12, pp. 3411–3419, 2019, doi: 10.1109/TBME.2019.2904885.

