## Supplementary material for "Patient-Specific Haemodynamic Analysis of Virtual Grafting Strategies in Type-B Aortic Dissection: Impact of Compliance Mismatch": Manuscript

A description of the mesh sensitivity study can be found in this supplementary material. The quality of the mesh and the analysis were assessed on six planes (Fig 1) of interest using the following metrics $f_i$: mean and maximum velocity and time average wall shear stress (TAWSS). Tin addition, therelative error between the metrics was computed between the coarse (M1) and medium (M2), and M2 and the fine (M3) meshes. Also, the grid convergence index (GCI) was computed following the study of Craven et al., [1], and the GCI was calculated as follows:

$r \sim (\frac{N_3}{N_2})^{1/3} \sim (\frac{N_3}{N_2})^{1/3}$w

$$p = \frac{\ln\left(\frac{|f_1 - f_2|}{|f_2 - f_3|}\right.}{\ln(r)}$$

$$E_{2,1} = \frac{|f_1 - f_2|}{f_2.(r^p - 1)} \qquad E_{3,2} = \frac{|f_2 - f_3|}{f_3.(r^p - 1)}$$

$$GCI_{2,1} = F_s|E_2| \qquad GCI_{3,2} = F_s|E_3|$$

With $N_{1,2,3}$ the number of elements of M1, M2 and M3, $f_{1,2,3}$ is the evaluated metric for each mesh, $F_S$ is a safety factor equal to 1.25 defined by Celik et al., [2] and used by Armour et al.,[3].

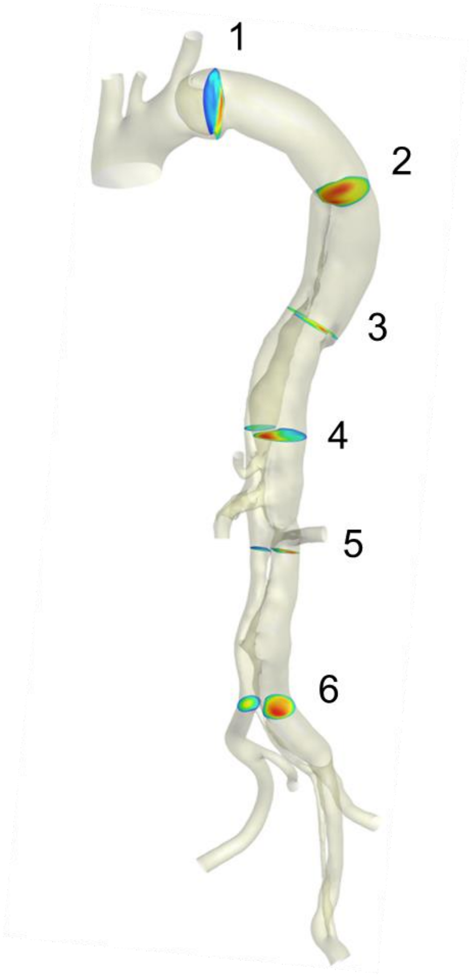


**Fig 1 Planes used for the mesh independence study**

Tables gathering all the measurements and derived calculus can be found bellow:

| Mean TAWSS | | | | | | |
|---|---|---|---|---|---|---|
| Plane | 1 | 2 | 3 | 4 | 5 | 6 |
| M1 [Pa] | 0.135 | 0.178 | 0.129 | 0.115 | 0.154 | 0.145 |
| M2 [Pa] | 0.136 | 0.185 | 0.130 | 0.116 | 0.156 | 0.144 |
| M3 [Pa] | 0.139 | 0.183 | 0.130 | 0.116 | 0.156 | 0.144 |
| $\%_{m,c}$ | 1% | 4% | 1% | 1% | 2% | -1% |
| $\%_{f,m}$ | 2% | -1% | 0% | 0% | 0% | 0% |
| $GCI_{2,1}$ | 0.41 | 7.29 | 6.94 | 2.60 | 3.03 | 1.50 |
| $GCI_{3,2}$ | 0.02 | 1.64 | 3.54 | 0.64 | 0.77 | 0.37 |

**Table 1 Mean TAWSS on the wall nodes of the planes of interest. Relative error and GCI between the specified mesh**

| Mean velocity | | | | | | |
|---|---|---|---|---|---|---|
| Plane | 1 | 2 | 3 | 4 | 5 | 6 |
| M1 [m/s] | 0.614 | 0.685 | 0.612 | 0.427 | 0.370 | 0.313 |
| M2 [m/s] | 0.651 | 0.845 | 0.583 | 0.455 | 0.346 | 0.302 |
| M3 [m/s] | 0.653 | 0.863 | 0.598 | 0.462 | 0.352 | 0.299 |
| $\%_{m,c}$ | 6% | 23% | -5% | 7% | -7% | -4% |
| $\%_{f,m}$ | 0% | 2% | 3% | 2% | 2% | -1% |
| $GCI_{2,1}$ | -2.41 | 2.26 | 0.78 | 1.88 | 0.75 | 1.56 |
| $GCI_{3,2}$ | -4.35 | 0.79 | 0.30 | 1.34 | 0.20 | 1.04 |

**Table 2 Mean velocity on the planes of interest. Relative error and GCI between the specified mesh**

| Maximum velocity | | | | | | |
|---|---|---|---|---|---|---|
| Plane | 1 | 2 | 3 | 4 | 5 | 6 |
| M1 [m/s] | 1.209 | 0.5409 | 0.4775 | 0.4221 | 0.9172 | 0.6162 |
| M2 [m/s] | 1.228 | 0.5903 | 0.479 | 0.4255 | 0.9283 | 0.6232 |
| M3 [m/s] | 1.238 | 0.5811 | 0.486 | 0.431 | 0.933 | 0.6239 |
| $\%_{m,c}$ | 2% | 9% | 0% | 1% | 1% | 1% |
| $\%_{f,m}$ | 1% | -2% | 1% | 1% | 1% | 0% |
| $GCI_{2,1}$ | 2.15 | 2.39 | -0.50 | -2.62 | 1.10 | 0.16 |
| $GCI_{3,2}$ | 1.12 | 0.45 | -2.29 | -4.18 | 0.46 | 0.02 |

**Table 3 Maximum velocity on the planes of interest. Relative error and GCI between the specified mesh**

[1] B. A. Craven, E. G. Paterson, G. S. Settles, and M. J. Lawson, “Development and verification of a high-fidelity computational fluid dynamics model of canine nasal airflow,” *J Biomech Eng*, vol. 131, no. 9, pp. 1–11, 2009, doi: 10.1115/1.3148202.

[2] I. B. Celik, U. Ghia, P. J. Roache, C. J. Freitas, H. Coleman, and P. E. Raad, “Procedure for estimation and reporting of uncertainty due to discretization in CFD applications,” *Journal of Fluids Engineering, Transactions of the ASME*, vol. 130, no. 7, pp. 0780011–0780014, 2008, doi: 10.1115/1.2960953.

[3] C. H. Armour *et al.*, “The influence of inlet velocity profile on predicted flow in type B aortic dissection,” *Biomech Model Mechanobiol*, vol. 20, no. 2, pp. 481–490, Apr. 2021, doi: 10.1007/s10237-020-01395-4.
